# Environmental and behavioral exposure pathways associated with diarrhea and enteric pathogen detection in twenty-six week old urban Kenyan infants: a cross-sectional study

**DOI:** 10.1101/2021.11.13.21266307

**Authors:** Kelly K. Baker, Jane Mumma, Sheillah Simiyu, Daniel Sewell, Kevin Tsai, John Anderson, Amy MacDougall, Robert Dreibelbis, Oliver Cumming

**Affiliations:** Department of Occupational and Environmental Health, University of Iowa, Iowa City, Iowa, United States; Center of Research, Great Lakes University of Kisumu, Kisumu, Kenya; Urbanization and Wellbeing unit, African Population and Health Research Center, Nairobi, Kenya; Department of Biostatistics, University of Iowa, Iowa City, Iowa, United States; Independent Research Consultant, Snowmass, Colorado, United States; Department of Disease Control, London School of Hygiene and Tropical Medicine, London, United Kingdom

## Abstract

The prevalence of enteric pathogen detection in children in low-income countries climbs rapidly between birth and 6 months of age. Few studies have tested whether improved household environmental and behavioral hygiene conditions protects infants from exposure to enteric pathogens spread via unhygienic human and animal sanitation conditions, especially during this early window of infancy. This cross-sectional study utilized enrollment survey data among households with 6 month old infants in Kisumu, Kenya participating in the Safe Start cluster-randomized controlled trial to estimate associations between household water access and treatment, animal vectors, sanitation access, hand washing practices, supplemental feeding, and flooring, with the outcomes of caregiver-reported 7-day diarrhea prevalence and sum count of different enteric viruses, bacteria, and parasites pathogens in infant stool. Then, we tested whether household environmental hygiene and behavioral practices moderated associations between infant exposure outcomes and latrine access and domestic animal co-habitation. We found that reported handwashing after handling animals and before eating were strongly associated with lower risk of caregiver-reported diarrhea, while owning and co-habitating with animals (versus no animals), living in a household with vinyl covered dirt floors (versus finished floors), and feeding infants cow milk (versus no milk) were strongly associated with pathogen detection in infants. Caregiver handwashing after child or self-defecation moderated the relationship between shared sanitation (vs private) sanitation access and infant exposure to pathogens such that handwashing had the greatest benefit for preventing pathogen exposure of infants in households with private latrines. In the absence of handwashing, access to private sanitation posed no benefits over shared latrines for protecting infants from exposure. Our evidence highlights eliminating animal co-habitation, improving flooring, improving post-defecation and food-related handwashing, and improving safety of cow milk sources and/or safe household storage of milk as interventions to prevent enteric pathogen exposure of infants less than 6 months age.

**Key Questions:** *What is already known?:* The population prevalence of enteric infections and diarrhea climbs rapidly in the first year of life. Risk factors for pediatric infections include unhygienic human and animal sanitary conditions that introduce feces into the environment, as well as intermediate environmental and behavioral exposure pathways. Research examining the mitigating role of improved environmental and behavioral conditions in preventing infant exposure to human and animal sanitary conditions is limited.

*What are the new findings?:* Contact with domestic animals and feeding infants cow milk are leading risk factors for exposure to enteric pathogens by 6 months age in Kisumu, while handwashing after animal handling and before eating are protective factors against self-reported diarrhea. The benefits of access to a private improved latrine (versus shared) for protecting infants from pathogen exposure were conditional upon caregivers washing hands after defecation or child-defecation.

*What do the new findings imply?:* Interventions that keep animals out of infant living spaces and that improve food-related and post-defecation handwashing may be the most effective strategies for controlling the population prevalence of enteric infections in infants between birth and 6 months age in Kisumu and similar settings.

## Introduction

More than a third of all children in low- and middle-income countries (LMICs) experience infections by one or more pathogens within the first year of life ^1,2^. Longitudinal studies of enteric pathogen prevalence show that the proportion of children in low-income communities shedding pathogens climbs rapidly after birth and is sustained through at least 24 months of age ^2,3^. One enteric infection episode can increase susceptibility to re- or co-infection through pathogen-pathogen interactions in the gut, downregulation of protective immune- or microbiome responses ^4^, and enteric enteropathy of intestinal tissue ^3,5^. Infants who experience early onset of infections (e.g. < 6 months of age) may be more vulnerable to early childhood infections.

Multiple interrelated environmental exposure pathways can contribute to pathogen transmission. Literature examining exposure pathways for infants in the first six months of life, and the effectiveness of interventions that target those pathways, beyond breastfeeding, is scarce. Evidence on impact of improving household safe drinking water sources and/or treatment and storage, access to basic latrines, and handwashing with soap on overall enteric pathogen detection and diarrhea in children under two years of age is mixed, with some rigorously controlled and well-powered trials of interventions reporting little impact, even with high levels of behavioral compliance ^1,6,7^. One explanation for the trial results is that interventions did not target the most important conditions resulting in pathogen transmission. Most hand washing interventions focus on infant caregivers, not on < 5 year siblings and playmates ^8^. Infant food safety depends upon hand washing, but also hygienic conditions of the food preparation and feeding environment ^9-11^. Additionally, urbanization has led to a rise in reliance on ready-to-eat roadside and packaged foods, which are rarely assessed for microbial safety. The presence of rodents, flies, and domestic animals in the immediate household or compound can contaminate soil and surfaces in areas where infants dwell with feces containing pathogens ^12-14^, and those pathogens could persist longer in the environment if household flooring is dirt or another type of permeable material ^15^. Similarly, the presence of older siblings who could transmit pathogens through child-child interaction or open defecation on floors could contaminate surfaces ^16^. The benefits of maintaining hygiene conditions in one’s own household may be derailed if hygiene conditions of communal compound living spaces and infrastructure (e.g. shared latrines) are poor ^17-21^. Access to a latrine within in a compound is neither a guarantee that all compound residents can consistently use it nor a guarantee that one’s neighbors will use it for child feces disposal even if it is accessible ^21^. Sharing latrines is a risk factor for childhood diarrhea^20^. At a larger scale, maintaining hygiene in one’s household may have little benefit if caregivers take infants to daycares or heavily polluted community settings, or if pollution from community is blown, flooded, or transported by foot traffic into compound grounds ^22-24 24^.

The goal of this study was to examine the role of intermediate household environmental (e.g. flooring, refrigeration) and behavioral (e.g. handwashing, water treatment) conditions in modifying pathogen exposure risks from human and animal feces sources for 6 month old infants in Kisumu, Kenya. This study utilizes enrollment data for infants in the Safe Start Trial^25^, a cluster-randomized trial of an infant food hygiene behavior change intervention. To achieve this goal, we estimated associations between water, animal, sanitation, hygiene, food, and flooring conditions with caregiver report of 7-day diarrhea prevalence in infants, and with sum count of pathogen types detected in infant stool as determined by molecular diagnostic assays. We then conducted a moderation analysis to examine a priori specified hypotheses of proposed environmental conditions and human behaviors that could influence the size and strength of association between feces sources and enteric pathogens in infants.

## Materials and Methods

### 2.1. Study Setting/Ethical Consideration

This cross-sectional study uses baseline data collected from six month old infants and their caregivers at the point of enrollment into Safe Start cluster-randomized controlled trial of an infant food hygiene behavior change intervention in Kisumu, Kenya (Clinical Trials identifier: NCT03468114). The formative work and trial protocol, including the estimation of sample size for evaluating trial impact, are described elsewhere ^25-29^. Kisumu is a city of approximately 490,000 people (Kisumu county integrated development plan 2013-2017) in the western region of Kenya. The study site includes communities in two low-income peri-urban neighborhoods in Kisumu. The study was approved by the scientific and ethical review committees at the GLUK (Ref. No. GREC/010/248/2016), LSHTM (Ref. No. 14695), and UI (IRB ID 201804204).

### 2.2. Study Design and Participants

Caregivers with six-month-old infants living in the catchment area of participating Community Health Volunteers (CHVs) were enrolled into the trial and participated in baseline data collection involving a survey and stool collection for microbial analysis. We defined eligibility of an infant as being 22 weeks (+/− 1 week) of age as verified by birth registration card who resided in one of the two study neighborhoods. We enrolled caregivers who were responsible for care of the infant during the day and were at least 18 years of age.

### 2.3. Outcomes

The outcomes for this study are: (1) 7-day caregiver-reported diarrhea prevalence prior to enrollment, where diarrhea is defined as three or more loose, watery stools in the previous 24 hours; and (2) the sum count of enteric pathogens detected in infant stool.

### 2.4. Data and Sample Collection

The study was described in the caregiver’s natural language, and a signed copy of the consent form was left for their records. Upon verification of eligibility and consent, caregivers were interviewed to collect data about household socio-economic conditions, access to water and sanitation infrastructure, animal ownership, and hygiene practices. In anticipation that infant breastfeeding and feeding practices may vary day to day and may be subject to response bias or recall bias, we asked caregivers about overall dietary history and foods given to infants in the last day. Caregivers were then provided several sterile commercially produced diapers and a sterile Ziploc bag, and asked to use these until the infant defecated. Diapers prevented cross-contamination via, for example, caregivers collecting infant feces with dirt attached from the ground. Caregivers were instructed to fold the diaper, place it into the storage bag, and store it in a cool dark place out of the reach of children and animals. The research team returned within 24 hours to collect the diaper, placed it in a cooler on ice packs, and transported it to the laboratory within five hours of collection. If the infant did not defecate in the first 24 hours, the team returned each day up to five days after enrollment to assess whether the infant had defecated. If no feces could be collected, the infant was de-enrolled from the study.

### 2.5. Nucleic Acid Extraction

Lab technicians unwrapped diapers in biosafety cabinets and used sterile stool collection scoops to transfer 200 mg of stool into Zymobiomics Shield Collection tubes, which were vortexed on a bead beater for 20 minutes and then processed according to the manufacturer’s instructions for the ZymoBIOMICS™ DNA/RNA extraction mini-kit (Zymo Research, Irvine, CA). One molecular-grade water only sample was prepared each day of sample processing as a process contamination control. Approximately half (N=383) of samples were spiked with 3 µL of 1.8*10^6^ CFU/µL of live bacteriophage MS2 to serve as a process control to assess for inhibition and efficiency in DNA and RNA recovery. Samples were transported on dry ice to the University of Iowa and stored at -80? until analysis.

### 2.6. TaqMan Array Card Analysis

A total of 23 gene targets of pathogen of interest in the TaqMan assays were used to assess pathogen presence in infants. Pathogen gene targets were: Adenovirus 40–41 Fiber, Adenovirus broad species Hexon, Rotavirus NSP3, Norovirus GI ORF 1-2, Norovirus GII ORF 1-2, Aeromonas aerolysin toxin *aerA, Campylobacter jejuni/C. coli (cadF), Enterohemorrhagic E. coli (EHEC) 0157 rdb, Enteroaggregative E. coli (EAEC) aatA and aaiC, Enteropathogenic E. coli (EPEC) bfpA and eae, Enterotoxigenic E. coli (ETEC) elt and est, Clostridium difficile tcdB, Salmonella enterica ttr, Shigella spp. virG, Vibrio cholerae hlyA, Giardia duodenalis* Assemblage A triosephosphate isomerase (TPI), Giardia duodenalis Assemblage A triosephosphate isomerase (TPI), *Cryptosporidium spp*. 18S, *C. hominus* LIB13, and *C. parvum* LIB13 ^30,31^. For each sample, 40 µL of extract was mixed with 5 µL of nucleic acid-free water, 50 µL of 2X RT-buffer, 0.6 ul of 50 mg/mL Bovine Serum Albumin (to reduce inhibition), and 4 µL of 25X AgPath enzyme from the Ag-Path-ID One-Step RT-PCR kit (Thermo Fisher, Waltham, MA) and pipetted into a well on a compartmentalized TaqMan card that included primer and probe assays in duplicate for each gene. TaqMan assays were completed in either a ViiA7 or QuantStudio8 instrument (Thermo Fisher, Waltham, MA), for cycling conditions: 45°C for 20 minutes and 95°C for 10 minutes, followed by 40 cycles of 95°C for 15 seconds and 60°C for 1 minute. A subset of samples that included both low and high Cq results were analyzed on both machines and compared to confirm that results did not vary between machines, before proceeding with further use of both machines. We defined a sample as positive if a gene target amplified within a 35 Cycle threshold (Cq35). If multiple gene targets were used to detect one type of pathogen, amplification of either gene (*EAEC aaiC/aatA, EPEC bfpA/eae, ETEC elt/est*) was considered positive for the general type of pathogen. The pathogen-specific detection patterns used to define the pathogen count variable for this analysis are described elsewhere.

### 2.7. Data Analysis

Independent variables representing point sources of feces that could contaminate the environment with enteric pathogens included household latrine design and location of the latrine, sharing of a latrine, ownership of domestic animals, whether animals are kept inside the household, and observation of rodents or their feces in the household. Improved sanitation was defined as a flush, pour flush, ventilated pit latrine, or pit latrine with an impermeable slab, according to WHO/UNICEF JMP criteria^32^. Independent variables representing environmental conditions or behaviors that are intermediate pathogen exposure pathways included: household flooring (soil/surfaces), primary and secondary water sources, whether the primary water source is intermittent, treating drinking water after collection, prior and recent (in the last day) breastfeeding status, prior and recent history of feeding the infant water or liquids other than breastmilk, history of feeding the infant solid foods, presence of a handwashing station with soap and water, and self-reported handwashing at critical times. A basic water source was defined as a piped tap to household or compound, a public tap, tube well, borehole, protected spring, protected hand dug well, or rainwater, according to World Health Organization (WHO)/UNICEF Joint Monitoring Programme criteria, available within 30 minutes round trip ^32^. Food ingredients, especially animal-based ones, can also be sources of pathogens that originate outside the household, but are treated here as exposure variables due to foods also being handled within the household before feeding the infant.

Potential confounders included in this analysis are marital status of caregiver, maternal education, household wealth, presence of multiple children under five years of age, infant preterm birth status, rotavirus vaccination status, and prior or current breastfeeding practices. Principal components analysis with Promax rotation of 15 household assets (bicycle, motorbike, car, refrigerator, mobile phone, wrist or pocket watch, wall clock, radio, cassette or CD player, television, DVD player, microwave oven, presence of grates on the windows and doors, use of electricity for lighting, and use of propane or electricity for cooking) resulted in a household wealth variable which was stratified into five quintiles.

All analysis were conducted with R software version 4.0.3 (R Foundation for Statistical Computing, Vienna, Austria). Associations between exposures and the binary indicator for 7-day caregiver-reported diarrhea were evaluated using logistic regression. Bivariate associations were evaluated between each exposure and self-reported diarrhea, and fully adjusted relationships between the exposures and the outcome was evaluated by running a single model with the confounder variables listed above and all exposure variables included. Associations between exposures and the ordinal categorical pathogen count variable were estimated using ordinal logistic regression. As with caregiver-reported diarrhea, we evaluated bivariate and fully adjusted relationships.

In both fully adjusted models, multicollinearity among the exposure variables resulted in non-identifiable or nearly-non-identifiable model, resulting in the exclusion of some redundant exposure variables. In addition, some confounder variables had insufficient variability to have estimable effects and hence were removed. Finally, caregivers reporting a lack of breastfeeding were so rare that they were excluded from the data. Logistic regression results are reported as odds ratios (OR), with 95% Confidence Intervals (95%CI), for having diarrhea, and ordinal logistic regression results are reported as ORs for having a higher count of pathogen types in stool versus fewer pathogen types. A random effect was initially included in models to adjust for spatial clustering, but was removed due to lack of variation in outcomes between villages.

Our moderation analyses tested whether hygiene of intermediate exposure pathways modified the relationships between point sources of pathogen contamination and infant health outcomes.

#### Hypothesis 1

Latrines could result in infant exposure to pathogens in human feces ^20^ through feces being tracked by feet onto household floors where infants play and place objects or hands that have been on the floor in their mouth. We hypothesize that floor type moderates the effect of latrine access on pathogens in infants such that permeable dirt floors that absorb liquids and sustain microbial growth, and that are harder to clean and disinfect, increase the association between human sanitation and pathogens in children compared to impermeable floors, like vinyl, concrete, or tile.

#### Hypothesis 2

Human sanitation can result in infant exposure to pathogens when caregivers do not wash hands after self-defecation or cleaning a child and then place hands in the infant’s mouth. We hypothesize that handwashing after defecation or child defecation moderates the effect of latrine access on infant health, such that human sanitation will be associated with pathogens in children among caregivers not washing hands after self or child defecation but not among caregivers who wash hands after self/child defecation.

#### Hypothesis 3

Like with latrines, associations between domestic animals or rodents and enteric infections in children could be caused by exposure of infants to floors contaminated with animal feces ^14^. We hypothesize that floor type can moderate this risk such that a dirt floor increases the association between domestic animals kept in or near the household or the presence of rodents and pathogens in children compared to households with impermeable floors.

#### Hypothesis 4

Zoonotic transmission of pathogens to infants could also occur through hands of caregivers who touch domestic animals or their feces, and then place hands in the infant mouth. We hypothesize that washing hands after handling animals moderates the effect of animal or rodent presence in the household on pathogens in children such that caregivers not washing hands after handling animals increases the association between animals or rodents and pathogens in infants compared to caregivers who wash hands after touching animals.

#### Hypothesis 5

Animal or human feces contamination on hands can be introduced into infant food during preparation or feeding and ingested by the infant while handwashing after defecation and animal handling could prevent transmission. We hypothesize that food-related handwashing also moderates the association between feces sources (latrine access and the presence of animals or rodents in the household) and pathogens in infants such that caregivers washing hands before preparing food and eating or feeding the infant decreases the association between latrines, domestic animals, and rodents with pathogens in infants, compared to caregivers who do not wash hands at these times.

#### Hypothesis 6

Infant supplemental foods include street foods prepared by vendors, raw fruits, and pre-packaged commercial products (e.g. pasteurized milk) that could contain contamination. Cooking (e.g. porridge), washing (e.g. fruit), or storing these foods can mitigate or enhance these external food system-based pathogen transmission risks ^33^. We hypothesize that access to a refrigerator for food storage moderates the pathway between supplemental foods and pathogens in infants such that a lack of refrigeration increases the association between supplemental food and pathogens in infants compared to households with refrigeration.

#### Hypothesis 7

Similarly, reliance on unsafe water sources can increase the chances of pathogen infection through water, but water treatment can reduce or eliminate this contamination. We hypothesize that filtering, boiling, or chlorinating water after collection moderates the association between type of water source and pathogens in infants such that not treating drinking water increases the association between household drinking water source and pathogens in infants compared to households who treat their drinking water.

These hypotheses were tested one at a time by adding an interaction term to the fully adjusted pathogen count model; the large number of potential confounders and exposure variables prohibited the simultaneous inclusion of all interaction terms being estimable. For each hypothesis listed above we ran an ordinal logistic regression model of the form

*# Pathogens = Confounder variables + Exposure variables + Hypothesis-specific interaction term*.

Moderation effects were then tested using a likelihood ratio test comparing the additive model excluding the interaction term to the full model including the interaction term.

## Results

### Characteristics of study participants

A total of 898 infant-caregiver pairs were enrolled in the study and completed the enrollment survey. The majority of caregivers were married, with secondary-level education, and had multiple young children (Table 1). The preterm birth rate was high at 17% of infants. Most infants had received two doses of oral rotavirus vaccine, as verified by infant health registration cards. Access to a basic drinking water source and a latrine of improved design was common, although a third of caretakers used secondary water sources and nearly all households shared their latrine with multiple other families. Most households had cleanable (ceramic, tile, concrete, or wood) floors or covered a dirt floor with rugs or vinyl. Of the 13.9% of households who owned animals, 86% (11.7% of all caregivers) kept their animals inside the household. Almost all caregivers reported that the infant was still breastfed, but only half reported giving the infant breastmilk in the last 24 hours. Almost a third of infants consumed cow or goat milk by six months of age, with Long Life ultra-high temperature (UHT) pasteurized milk being the most common supplemental food. In the last day prior to the interview, a fifth of infants were given water and a third cow’s milk, with UHT being the most common. Only ∼5% of caregivers reported feeding infants solid foods. A handwashing station with soap and water was observed in few (6.8%) households, yet handwashing at critical times was reported by over 60% of caregivers for after self-defecation or infant defecation, and for preparing food, feeding the infant, and personal eating. In contrast, just 12.8% reported washing hands after handing animals.

**Table 1.** Characteristics of caregivers and their 6-month infants enrolled into the Safe Start Study in Kisumu, Kenya between 2018 and 2019.

| **Socio-economic, environmental, and behavioral living conditions** |  |
| --- | --- |
| Caregiver is single vs Married | 11.8 (105) |
| Caregiver has primary or no education vs secondary or higher education | 39.1 (348) |
| Household wealth Quintile |  |
| - Poorest | 19.6 (174) |
| - Low-middle | 19.7 (175) |
| - Middle | 20.7 (184) |
| - Upper-middle | 20.0 (178) |
| - Highest quintile | 20.0 (178) |
| More than one child under five years lives in household | 35.6 (320) |
| Infant was born preterm (<37 weeks gestational age) vs not preterm (≥37 gestational weeks age) | 16.0 (140) |
| Rotavirus vaccination series <sup>1</sup> |  |
| - No vaccination | 1.2 (10) |
| - One dose | 1.6 (13) |
| - Both doses | 97.2 (803) |
| Infant ever breastfed vs never breastfed | 99.8 (885) |
| Infant still breastfed vs weaned | 99.7 (884) |
| Breastfed in the last day vs not breastfed in last day | 57.5 (516) |
| **Exposure variables** |  |
| Household relies on Improved latrine vs unimproved latrine or open defecation <sup>2</sup> | 86.6 (770) |
| Sharing latrine |  |
| - Private household latrine | 10.9 (98) |
| - 1-5 households share | 17.7 (159) |
| - >5 households Share latrine | 71.4 (641) |
| Household owns animals vs does not own animals | 12.8 (114) |
| Animals sleep inside household vs do not sleep in household | 10.5 (94) |
| Rodents or rodent droppings observed vs no droppings | 69.7 (620) |
| Household Flooring |  |
| - Unfinished dirt floor | 8.4 (75) |
| - Unfinished floor covered with vinyl sheets or carpet | 23.2 (206) |
| - Finished floor (ceramic, concrete, tile, wood) | 68.4 (608) |
| Household primarily relies on basic drinking water source vs limited or unimproved source <sup>2</sup> | 99.2 (885) |
| Household uses limited/unimproved secondary water source vs improved or no secondary source | 32.0 (287) |
| Household does not treat drinking water vs treats water by boiling, Water Guard, or Aqua Tabs | 43.8 (393) |
| Infant fed water in the last day vs did not give water | 18.8 (169) |
| Infant ever fed milk |  |
| - Never | 71.3 (632) |
| - Ultra heat treated (UHT) | 17.5 (155) |
| - Fresh packed milk | 8.9 (79) |
| - Local cow or goat | 1.0 (9) |
| - Powdered formula | 1.4 (12) |
| Infant has been fed solid food before vs never fed solid food | 4.7 (42) |
| Infant fed milk in the last day |  |
| - UHT vs no UHT | 11.2 (101) |
| - fresh packed vs no fresh packed milk | 2.0 (18) |
| - local cow milk vs no local milk | 0.8 (7) |
| - ATM Milk vs no ATM milk | 3.5 (31) |
| - Milk Bar vs no Milk Bar | 8.4 (75) |
| - Other milk-based liquid vs no other liquid | 4.0 (36) |
| Observed designated handwashing area with soap and water vs no handwashing area or no area with soap and water | 6.6 (59) |
| Reported Handwashing at critical times |  |
| - After cleaning an infant that defecated | 58.8 (528) |
| - After self-defecation | 88.0 (790) |
| - After handling animals | 12.0 (108) |
| - Before preparing food | 59.1 (531) |
| - Before feeding child | 60.5 (543) |
| - Before eating | 69.7 (626) |
| **Total enrolled infants** | **N=898** |

<sup>1</sup> Missing 64. <sup>2</sup> Defined according to WHO/UNICEF Joint Monitoring Programme criteria.

Variables excluded from multivariable exposure models due to low variation in sub-groups included rotavirus vaccination, infant ever breastfed, infant still breastfed, primary drinking water source, infant has been fed solid food before and ever feeding the infant Local Milk or Other milk type ever (UHT and fresh packed milk were only types with sufficient data). Breastfeeding the infant in the last day was used to control for confounding of diarrhea or pathogen detection by breastfeeding. Variables excluded due to collinearity included animals kept inside the household (nearly perfectly predicted by animal ownership) and feeding the infant milk in the last day (proxy for history of milk feeding).

### Exposure pathways and 7-day caregiver-reported diarrhea history

A total of 862 out of 898 caregivers interviewed at enrollment had complete data for all exposure variables of interest and self-reported 7-day diarrhea in infants. There were no discernible differences between individuals with and without full exposure data. Diarrhea prevalence among 6-month old infants in Kisumu at enrollment was 14.9% (134). Associations with potential confounders are reported in STable 1. In the full multivariable model, washing hands after handling animals and before eating were strongly associated with a 5-fold and 2.27-fold lower odds of reported diarrhea, respectively (Table 2). Use of improved latrines, washing hands after self-defecation, and washing before feeding infants was also protective against diarrhea, but these relationships were weakened after adjusting for other WASH conditions. Odds of diarrhea were higher for infants fed UHT and fresh packed milk, but these relationships were weakened after adjusting for other WASH conditions.

**Table 2.**
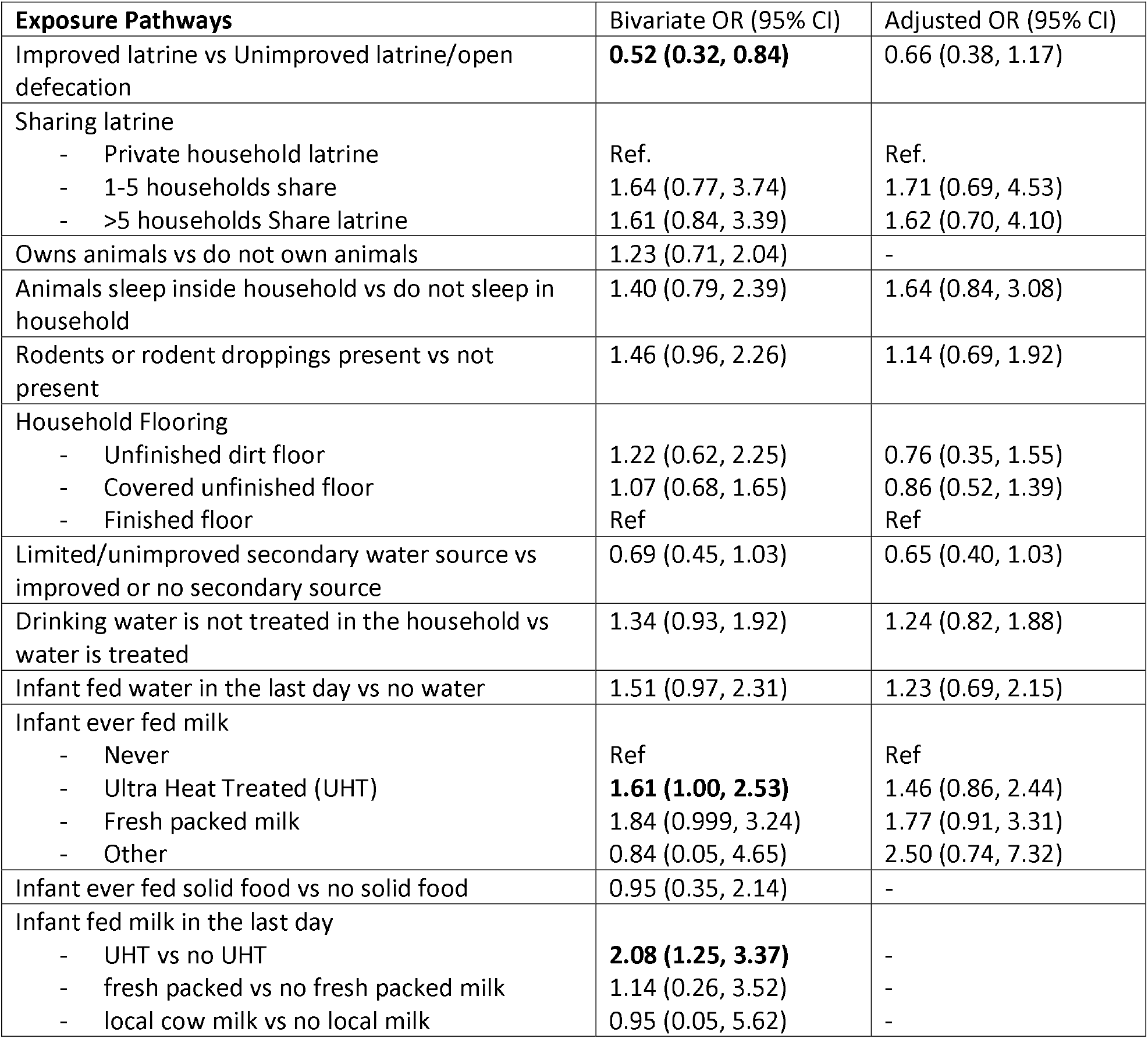
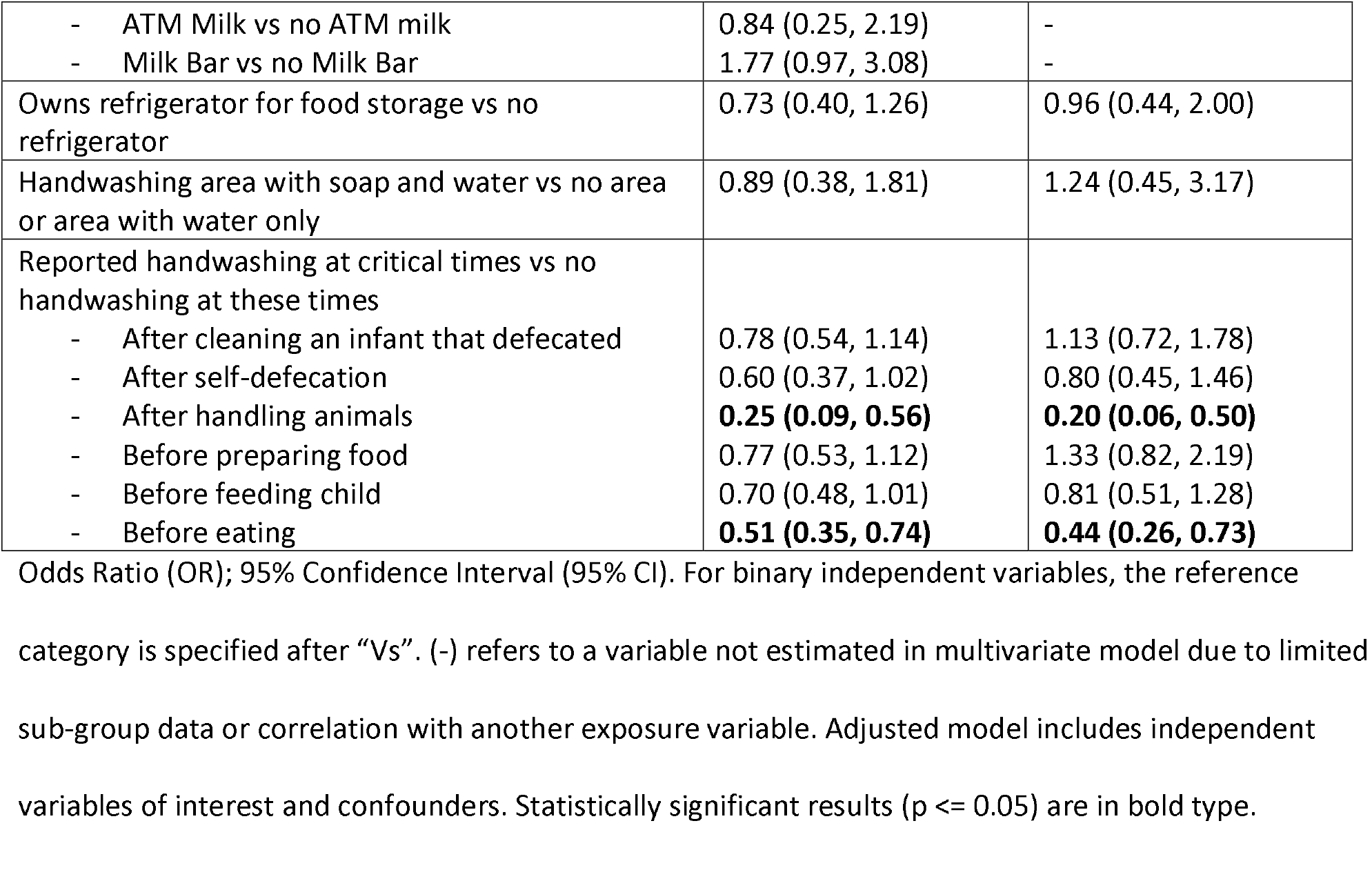
Odds ratios (OR) and confidence intervals (CI) from logistic regression models of caregiver-reported 7-day diarrhea in 6 month infants and potential feces sources and exposure pathways.

| **Exposure Pathways** | **Bivariate OR (95% CI)** | **Adjusted OR (95% CI)** |
| --- | --- | --- |
| Improved latrine vs Unimproved latrine/open defecation | **0.52 (0.32, 0.84)** | 0.66 (0.38, 1.17) |
| Sharing latrine |  |  |
| - Private household latrine | Ref. | Ref. |
| - 1-5 households share | 1.64 (0.77, 3.74) | 1.71 (0.69, 4.53) |
| - >5 households Share latrine | 1.61 (0.84, 3.39) | 1.62 (0.70, 4.10) |
| Owens animals vs do not own animals | 1.23 (0.71, 2.04) | - |
| Animals sleep inside household vs do not sleep in household | 1.40 (0.79, 2.39) | 1.64 (0.84, 3.08) |
| Rodents or rodent droppings present vs not present | 1.46 (0.96, 2.26) | 1.14 (0.69, 1.92) |
| Household Flooring |  |  |
| - Unfinished dirt floor | 1.22 (0.62, 2.25) | 0.76 (0.35, 1.55) |
| - Covered unfinished floor | 1.07 (0.68, 1.65) | 0.86 (0.52, 1.39) |
| - Finished floor | Ref | Ref |
| Limited/unimproved secondary water source vs improved or no secondary source | 0.69 (0.45, 1.03) | 0.65 (0.40, 1.03) |
| Drinking water is not treated in the household vs water is treated | 1.34 (0.93, 1.92) | 1.24 (0.82, 1.88) |
| Infant fed water in the last day vs no water | 1.51 (0.97, 2.31) | 1.23 (0.69, 2.15) |
| Infant ever fed milk |  |  |
| - Never | Ref | Ref |
| - Ultra Heat Treated (UHT) | **1.61 (1.00, 2.53)** | 1.46 (0.86, 2.44) |
| - Fresh packed milk | 1.84 (0.999, 3.24) | 1.77 (0.91, 3.31) |
| - Other | 0.84 (0.05, 4.65) | 2.50 (0.74, 7.32) |
| Infant ever fed solid food vs no solid food | 0.95 (0.35, 2.14) | - |
| Infant fed milk in the last day |  |  |
| - UHT vs no UHT | **2.08 (1.25, 3.37)** | - |
| - fresh packed vs no fresh packed milk | 1.14 (0.26, 3.52) | - |
| - local cow milk vs no local milk | 0.95 (0.05, 5.62) | - |
| - ATM Milk vs no ATM milk | 0.84 (0.25, 2.19) | - |
| - Milk Bar vs no Milk Bar | 1.77 (0.97, 3.08) | - |
| Owens refrigerator for food storage vs no refrigerator | 0.73 (0.40, 1.26) | 0.96 (0.44, 2.00) |
| Handwashing area with soap and water vs no area or area with water only | 0.89 (0.38, 1.81) | 1.24 (0.45, 3.17) |
| Reported handwashing at critical times vs no handwashing at these times |  |  |
| - After cleaning an infant that defecated | 0.78 (0.54, 1.14) | 1.13 (0.72, 1.78) |
| - After self-defecation | 0.60 (0.37, 1.02) | 0.80 (0.45, 1.46) |
| - After handling animals | **0.25 (0.09, 0.56)** | **0.20 (0.06, 0.50)** |
| - Before preparing food | 0.77 (0.53, 1.12) | 1.33 (0.82, 2.19) |
| - Before feeding child | 0.70 (0.48, 1.01) | 0.81 (0.51, 1.28) |
| - Before eating | **0.51 (0.35, 0.74)** | **0.44 (0.26, 0.73)** |

Odds Ratio (OR); 95% Confidence Interval (95% CI). For binary independent variables, the reference category is specified after “Vs”. (-) refers to a variable not estimated in multivariate model due to limited sub-group data or correlation with another exposure variable. Adjusted model includes independent variables of interest and confounders. Statistically significant results ( $p \leq 0.05$ ) are in bold type.

**Table 3.**
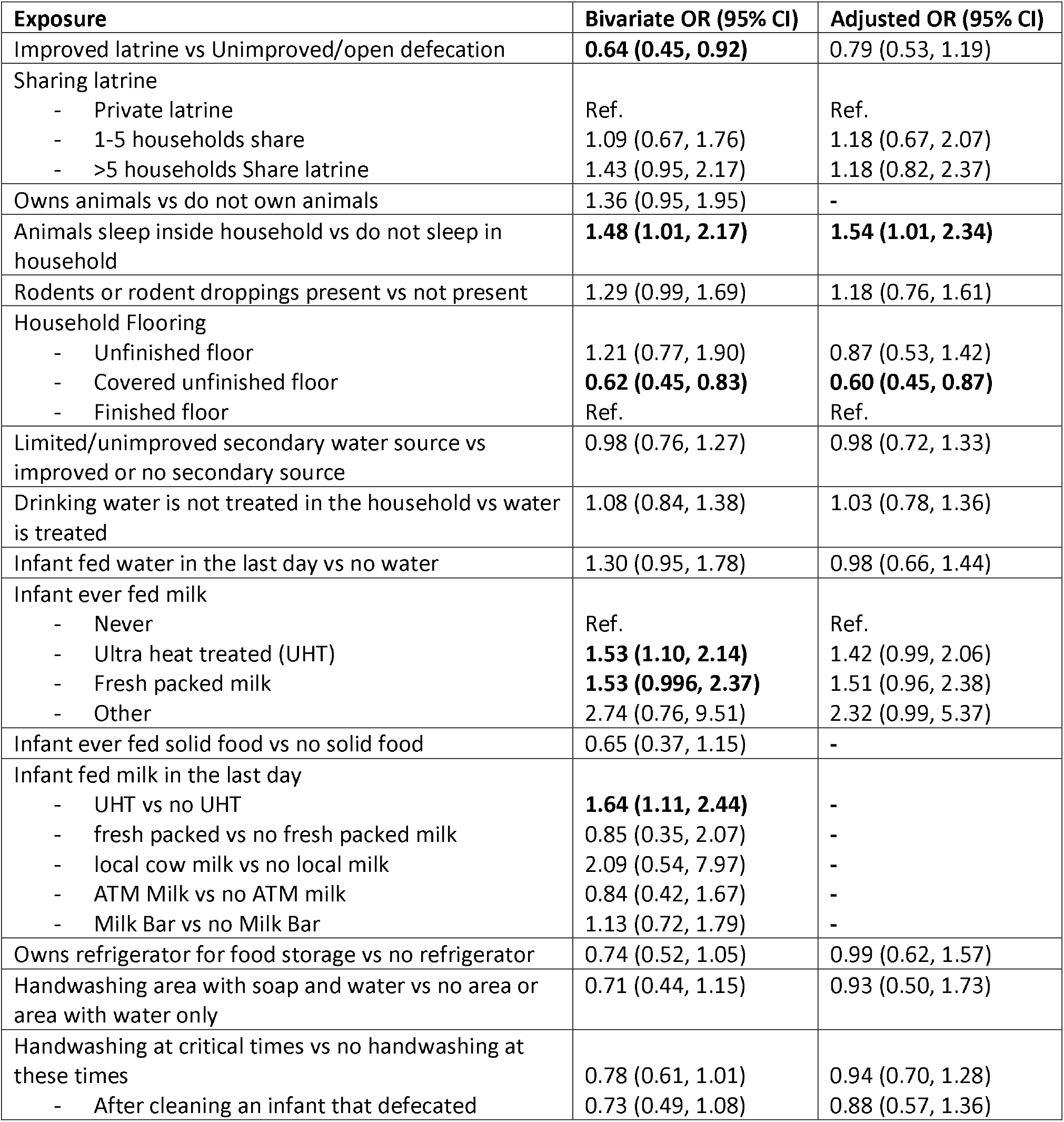
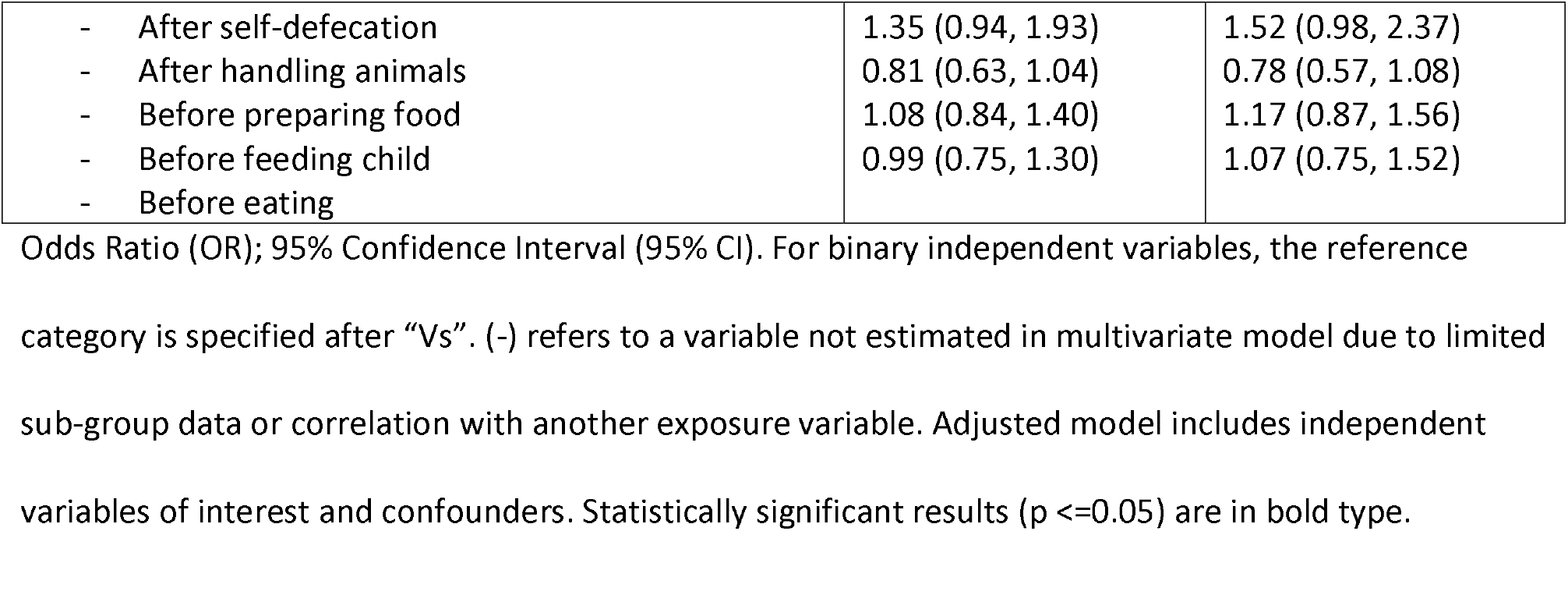
Odds ratios (OR) and confidence intervals (CI) from ordinal regression models of detecting a higher number versus lower number of enteric pathogen types in feces of 6 month infants and potential feces sources and exposure pathways.

### Exposure pathways and enteric pathogens in infants

Data on both the exposures of interest and pathogen detection outcome were available for 773 infants. Genes for at least one of 23 assessed enteric pathogens were detected in stool in 88.9% (n=707) of infants, with two or more pathogens detected in 66.9% (n=532) of infants (median 2 (25-75% Inter Quartile Range 1 - 3), based upon a Cq35 threshold. The ordinal categorical model outcome variable was categorized as one of 0, 1, 2, 3, 4, 5, or ≥6 types of pathogens detected, due to scarce data for higher pathogen counts. Associations with confounders are reported in STable 2. In the multivariable ordinal regression model, living in a household with a vinyl-covered dirt floor was associated with a 1.67 fold lower chance of having more pathogen types detected in infant stool compared to infants in households with cement or tile floors. Infants fed any type of milk had a 1.42 to 2.32 fold increased chance of a higher count in pathogen diversity in infant stool versus infants never given milk. Keeping animals in the house posed a 1.54 fold higher odds of higher pathogen count compared to not owning animals, while washing hands after handling animals was counterintuitively associated with a 1.52 fold increase in odds compared to not washing hands after handling animals.

### Moderation of Pathogen Exposure Pathways

After adjusting for confounders, moderation hypothesis tests for exposures and pathogen infections revealed:

1. There was no evidence that floor type modified the relationship between basic latrine access (p=0.93) or latrine sharing (p=0.49) and pathogen count in infants.
2. There was no evidence that handwashing after personal defecation (p=0.91) or after cleaning a child (p=0.96) modified the relationship between basic latrine access and pathogen count. There was strong evidence for the hypothesis that handwashing after personal defecation (p=0.03) or after child defecation (p=0.02) modified the relationship between latrine sharing and pathogen count (Figure 1A and 1B respectively). Sharing a latrine with 2 to 5 (OR=1.35; 95% CI: 0.75, 2.43) or >5 other households (OR=1.72; 95% CI: 0.99, 2.98) was associated with higher pathogen count in infants relative to households with private latrines among caregivers who washed hands post-defecation. Sharing a latrine with 2 to 5 (OR=1.79; 95% CI: 0.92, 3.51) or >5 households (OR=2.11; 95% CI: 1.13, 3.94) versus private latrines was also associated with higher pathogen detection in infants among caregivers who washed hands after child defecation. However, pathogen count was lower in households sharing a latrine with 2 to 5 (OR=0.26; 95% CI: 0.04, 1.62) or >5 other (OR=0.18; 95% CI: 0.04, 0.90) households among caregivers who did not wash after defecation, or after child defecation (OR=0.47; 95% CI: 0.18, 1.21; and OR=0.58; 95% CI: 0.24, 1.36, respectively). If sanitation is viewed as the effect modifier, then higher pathogen count was strongly associated with not washing hands after defecation (OR=8.43; 95% CI 1.71, 41.56) or child defecation (OR=3.59; 95% CI 1.35, 9.51) in households with private latrines. The association between pathogen count and not washing hands after defecation or after child defecation was weaker in households sharing latrines with 2-5 (OR=2.18; 95% CI 0.71, 6.72 and OR=1.67; 95% CI 0.78, 3.57, resp.) or >5 (OR=1.54; 95% CI 0.77, 3.09, and 2.06; 95% CI 1.07, 3.97, resp.) other households.
3. There was no evidence that floor type modified the relationship between owning domestic animals, and by proxy keeping them in the household (p=0.60) and pathogen count in infants.
4. There was no evidence that handwashing after handling animals modified the relationship between keeping animals inside and pathogen count (p=0.28).
5. There was no evidence that washing hands before food preparation modified the association between pathogen count and latrine access (p=0.74), sharing a latrine (p=0.49), or animals sleeping inside (p=0.24). Similarly, there was no evidence that washing hands before feeding a child modified the association between pathogen count and latrine access (p=0.54), sharing a latrine (p=0.61), or owning animals (p=0.94). And, there was no evidence that washing hands before self-eating modified the association between pathogen count and latrine access (p=0.15), sharing a latrine (p=0.47), or owning animals (p=0.63).
6. There was no evidence that access to household refrigeration modified the effect of milk type on pathogen count (p=0.17).
7. We could not investigate modification of treating water on unsafe primary water sources and pathogens, but among the 99.8% of households using a basic water source, there was no evidence that treating water by chlorination or boiling affected pathogen count in children (p=0.97).

**Figure 1.**
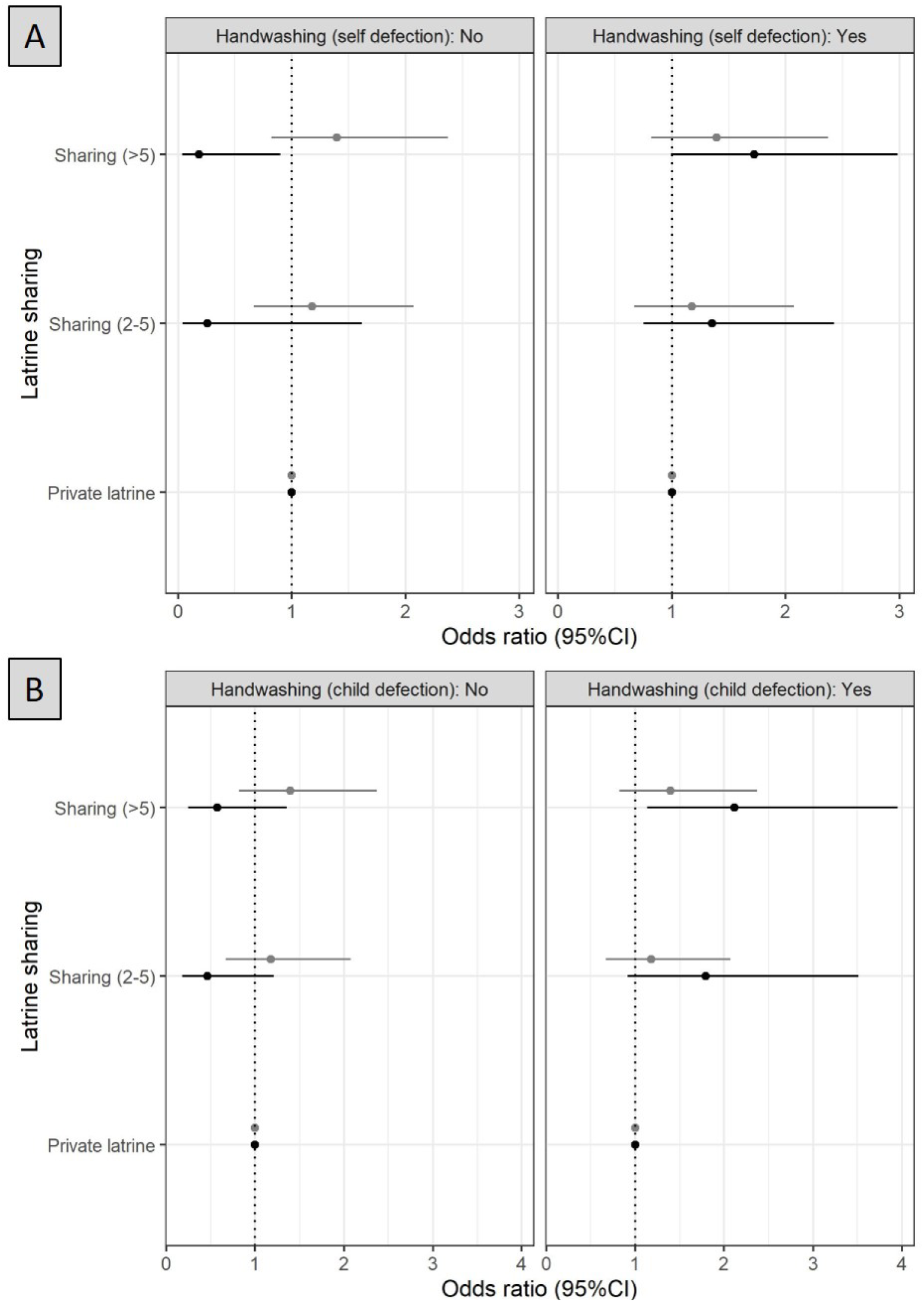
Effect modification of associations between shared and private sanitation and pathogen count in infants by handwashing after (A) self-defecation and (B) cleaning a child that defecated.

Figure 2 summarizes the relationships identified between human sanitation and animal presence conditions that could introduce pathogens into the household, the housing conditions that can transmit pathogens, and higher versus lower pathogen count in six month old infants.

**Figure 2.**
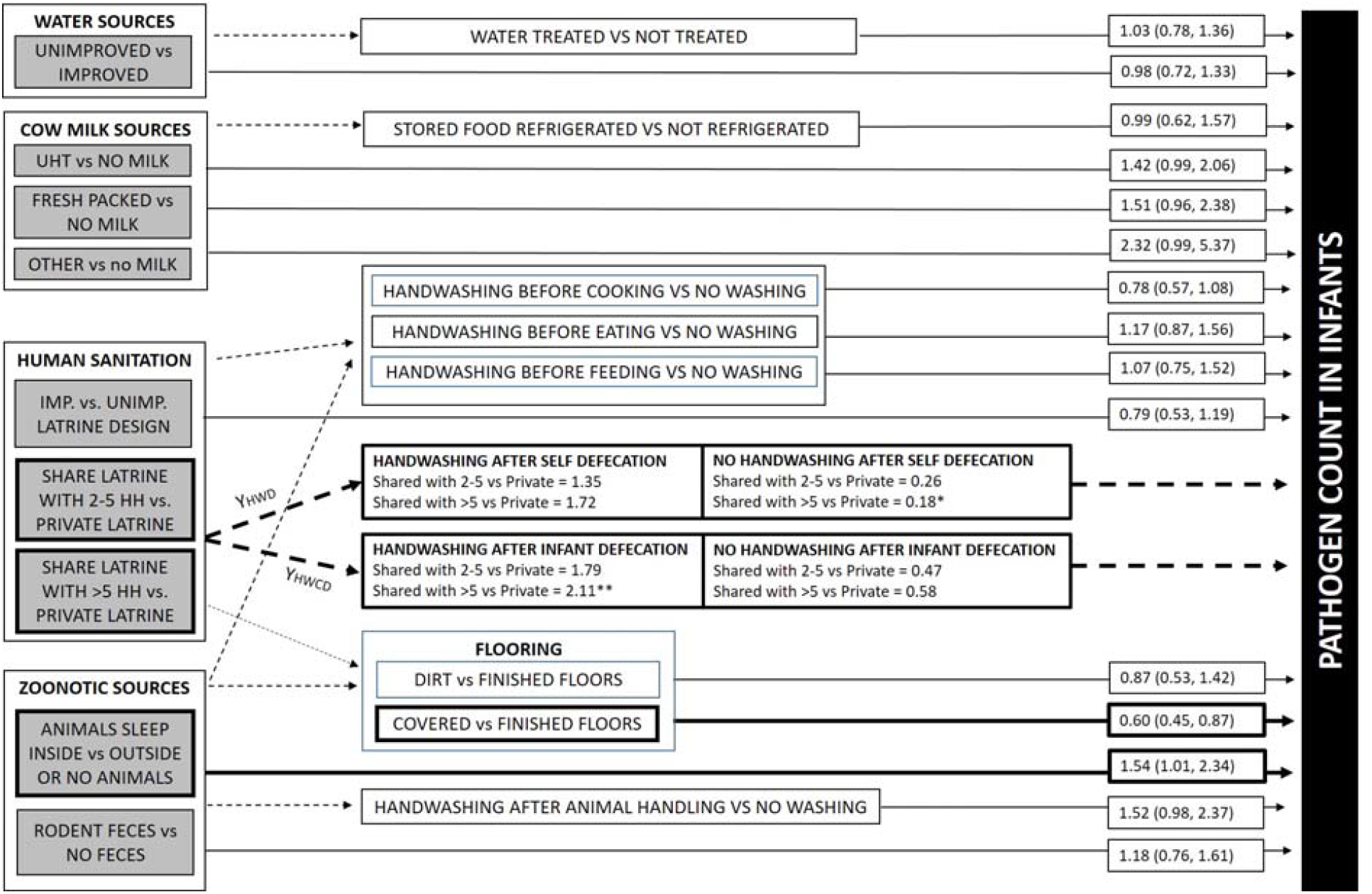
Size and strength of relationships between human and animal household sanitary conditions, potential food, soil/surface, and hand pathways that could transmit pathogens, and the odds of detecting higher versus lower counts in enteric pathogen types in infant feces. Footnote: Arrows representing each relationship are labeled with effect size and weighted in width and color based upon size of effect away from a null effect. Strength of effect is indicated by * 0.05<p<0.10; ** 0.01<p<0.05; and *** p<0.01. If effect modification of the pathogen source to infant outcome was identified, the source to modifier relationship is indicated by solid line (vs dotted for no evidence of modification) and conditional effects for pathogen source and outcome are reported by strata of the effect modifier. Some variables analyzed with minimal relationships to the outcome are omitted to reduce complexity of the image.

## Discussion

The goal of this study was to identify risk factors linked to diarrhea prevalence and enteric pathogen detection among < 6 month old infants in Kisumu, Kenya, and examine whether improved household hygiene environments (e.g. flooring, refrigeration for safe food storage) and behaviors (e.g. handwashing, water treatment) modified infant exposure to enteric pathogens from human and animal vectors. The very high prevalence of enteric pathogen detection and co-detection in Kisumu infants at 6 months age highlighted the critical need for exposure prevention interventions in early infancy, similar to other studies in low-income countries^2^. Our evidence implicated handwashing after handling animals and before eating and living in a household with vinyl covered dirt floors (vs finished floors) as strong factors reducing the risk of self-reported diarrhea at six months of age, while owning and co-habitating with animals and feeding infants cow milk were strong factors increasing the risk of exposure to pathogens. Additionally, our evidence suggested the benefits of private sanitation are limited if post-defecation handwashing is not practiced.

Latrine access was not a strong risk factor for diarrhea or pathogens in infants after adjusting for other environmental and behavioral conditions, although sharing latrines was implicated in pathogen exposure when examined in the context of mitigating post-defecation handwashing behaviors. The interaction between these two conditions was influenced by strong benefits from post-defecation handwashing in households with private sanitation, and sustained pathogen exposure risks to infants from shared latrines, even when caregivers were washing hands after touching feces. Infants usually have no direct contact with latrines designed for able-bodied children and adults, although shared latrines have been identified as a risk factor for moderate and severe diarrhea in children across countries and studies ^20,34^. Pathogen transmission from shared latrines could occur through pathways not examined in this study, like fly density, flooding, or soil on shoes or feet moving between the latrine and the compound.

Household flooring type has been minimally explored in enteric disease literature, but finished flooring has been linked to decreased risk from diarrhea in Egypt ^15^, and lower prevalence of *Ascaris lumbricoides* and soil-transmitted helminths in Kenya and Bangladesh in children under five years ^35^. In our study, we used household flooring construction as a proxy for floor hygiene, based upon the premise that covered or finished floors are easier to clean and less absorbent for sustaining pathogens. Type of household flooring did not modify the relationship between shared sanitation and pathogens in infants in this study. In fact, we counterintuitively observed higher pathogen counts in infants in households with the highest standard of finished flooring (mostly concrete) versus vinyl covered dirt floors. This could mean concrete, which is porous, can sustain pathogen contamination similar to dirt floors or that floor type was a poor indicator of floor hygiene. Caregivers who cover dirt floors with vinyl or carpet may have been more concerned about hygiene issues posed by living in undeveloped household structures, and clean them more often than caregivers who are not concerned about dirt floors or who have finished floors that they perceive to be safe. We did not ask caregivers about floor cleaning practices, which might be a better indicator on the role of floors in transmission.

Owning and sharing living spaces with animals was associated with higher pathogen counts. Co-habitation between infants and animals in domestic settings makes infant contact with animals or their feces likely, such as from ingestion of contaminated household drinking water ^36^. Additionally, domestic animals belonging to one’s neighbors often wander through spaces of households who do not own animals ^37^, meaning ownership is not a comprehensive indicator of animal exposure. While handwashing after animal handling was not associated with pathogen detection count and did not moderate the strong association between owning animals and pathogen count, it was protective against infant diarrhea. Owning animals may influence infant exposure via multiple pathways, with caregiver hands having relatively little importance as a pathway. We did not assess caregiver washing of infant hands, but infants place their own hands into their mouth frequently^23,38^. Touching of animals, animal feces, or contaminated floors, followed by placing hands, dirt, or feces in the mouth may be a more important mechanism linking animal presence and pathogens in infants.

Information on the role of supplemental foods in infant infection in the first months of life is also scarce, possibly due to the assumption that breastfeeding is the only or primary source of infant food at this stage. Like many self-reported behaviors where respondents are aware of sanctioned and disapproved behaviors, caregivers may over report breastfeeding to avoid censure. While nearly all caregivers said they still breastfeed 6 month infants, only 57% reported breastfeeding within the last day, reinforcing that supplemental foods are important exposure pathways for Kisumu infants younger than 6 months age. Those not breastfeeding relied mostly on cow’s milk, especially Long Life UHT milk. Urbanization of low-income cities has led to increased numbers of people living in low-income neighborhoods and more women working outside of the home. More women working outside the home has led to increased demand for convenient supplemental foods, like packaged milk, that are palatable to very young infants and can be used by secondary caregivers for feeding ^39-44^. Our data indicated packaged milk-based foods were sources of pathogen exposure, prompting the question as to whether the risks from UHT milk consumption derive from food supply chains or unhygienic household food management.

Animal-based foods, like cow milk, pose higher risks for enteric pathogen transmission than plant-based foods ^11^ although pasteurization is an effective means for making milk safer. In a parallel study, we showed enteric pathogens do occur in UHT milk products in Kenya, but households contribute more to infant food contamination than food products ^33^. Pasteurized milk products may be risk factors in this study due to post-purchasing contamination from household surfaces, hands, or utensils or storing milk for prolonged periods after opening. Food-related handwashing did not modify human and animal source-to-infant transmission, although washing before eating was generally protective for infants. Other practices related to the boiling, feeding, and storage of food were not studied here, but may explain or moderate foodborne pathogen exposure. Forthcoming manuscripts from the Safe Start trial will reveal whether promotion of handwashing during food preparation and feeding, boiling food, storing foods in closed containers, and feeding infants from dedicated containers can reduce diarrhea and enteric pathogen detection rates in this study population.

Additional limitations of the study include inability to assess the causal relationships between exposure conditions and outcomes, although it is unlikely the pathogen status of infants – to which caregivers were unaware – caused a change in household environments or behaviors. We also relied on self-reported response about breastfeeding, handwashing and feeding behaviors, and 7-day diarrhea symptoms in infants, which are all vulnerable to reporting bias. Enumerators visually verified as many conditions, such as water sources, latrines, handwashing station, and flooring as feasible. Additional strengths include the use of an ordinal distribution representing pathogens co-detection patterns for identifying many exposure risk factors that would have remained masked by using a simplistic pathogen presence/absence outcome. The utility of using microbial diversity in pathogen contamination for distinguishing between high and low risk conditions in settings where exposure is the norm has also been demonstrated in environmental studies of soil, water, and food ^22,26,45^. This study was performed among low-income peri-urban and urban Kenya households and the results may not be generalizable to rural households in Kenya, to higher-income households, and to low-income households in other countries.

In summary, our results point to a need for interventions that limit the presence of animals in households, increase handwashing after handling animals and using the latrine, and before eating, and promote safe management of milk-based infant foods to reduce the high population prevalence of enteric disease in < 6 month infants. Interventions must target commercial packaged cow milk as one of the most common supplemental foods, including packaged milk products that are rapidly growing in popularity in urban populations.

## Data Availability

All data produced in the present study are available upon reasonable request to the authors

## Author Contributions

KKB and OC conceived of this study concept. JM, OC, KKB, JA, and RD designed the parent study. SS and KT collected data. JM and KKB supervised data collection. JA managed data assimilation and curation. KKB and DS performed the analysis and drafted the manuscript. AM provided additional statistical review. All authors reviewed and submitted comments on the final manuscript.

## Acknowledgments

This study would not have been possible without the efforts of Rose Evalyn Aseyo, Edwin Atitwa, Bonphace Okoth, John Agira, Stephanie Houser, and an army of dedicated enumerators and Community Health Volunteers. The Safe Start trial, on which the study built, was funded by the United Kingdom Department for International Development (DFID) through the SHARE Research Consortium.

## License Statement A

I, the Submitting Author has the right to grant and does grant on behalf of all authors of the Work (as defined in the below author licence), an exclusive licence and/or a non-exclusive licence for contributions from authors who are: i) UK Crown employees; ii) where BMJ has agreed a CC-BY licence shall apply, and/or iii) in accordance with the terms applicable for US Federal Government officers or employees acting as part of their official duties; on a worldwide, perpetual, irrevocable, royalty-free basis to BMJ Publishing Group Ltd (“BMJ”) its licensees and where the relevant Journal is co-owned by BMJ to the co-owners of the Journal, to publish the Work in BMJ Global Health and any other BMJ products and to exploit all rights, as set out in our <u>licence</u>.

The Submitting Author accepts and understands that any supply made under these terms is made by BMJ to the Submitting Author unless you are acting as an employee on behalf of your employer or a postgraduate student of an affiliated institution which is paying any applicable article publishing charge (“APC”) for Open Access articles. Where the Submitting Author wishes to make the Work available on an Open Access basis (and intends to pay the relevant APC), the terms of reuse of such Open Access shall be governed by a Creative Commons licence – details of these licences and which <u>Creative Commons</u> licence will apply to this Work are set out in our licence referred to above.

