## Supplemental Materials for "Environmental and behavioral exposure pathways associated with diarrhea and enteric pathogen detection in twenty-six week old urban Kenyan infants: a cross-sectional study"

**Supplemental Figures**

**STable1.** Odds of caregiver-reported 7-day diarrhea in 26 week aged infants for confounders used to adjust the multivariable model.

| **Confounders** | **Bivariate OR (95% CI)** | **Adjusted OR (95% CI) ^1^** |
| --- | --- | --- |
| Single vs married | **1.82 (1.08, 2.95)** | 1.71 (0.95, 2.99) |
| Secondary or higher vs primary or no maternal education | **1.46 (1.01, 2.12)** | 1.40 (0.90, 2.18) |
| Wealth index quintile   - Highest - Above middle - Middle - Lower middle - Lowest | Ref.  1.40 (0.76, 2.59)  1.29 (0.70, 2.39)  1.42 (0.77, 2.64)  1.56 (0.86, 2.87) | Ref.  0.97 (0.47, 2.07)  0.80 (0.36, 1.81)  0.86 (0.38, 1.99)  0.89 (0.37, 2.16) |
| More than one child vs one child | 1.13 (0.77, 1.65) | 1.04 (0.68, 1.57) |
| Preterm versus term birth | **0.38 (0.18, 0.71)** | **0.43 (0.20, 0.85)** |
| Infant breastfed in the last day versus not breastfed | 0.96 (0.67, 1.40) | 0.77 (0.49, 1.22) |

Odds ratio (OR); Confidence Interval (CI).

**^1^** Adjusted for confounders and water, animal, sanitation, hygiene, food, and flooring variables of interest.

**STable 2.** Odds of detecting a higher number versus lower number of pathogen types in feces of 26 week aged infants for confounders used to adjust multivariable model.

| **Confounders** | **Bivariate OR (95% CI)** | **Adjusted OR (95% CI) ^1^** |
| --- | --- | --- |
| Single vs married | 0.97 (0.66, 1.42) | 0.84 (0.55, 1.28) |
| Secondary or higher vs primary or no maternal education | **1.31 (1.02, 1.69)** | 1.09 (0.82, 1.45) |
| Wealth index quintile   - Highest - Above middle - Middle - Lower middle - Lowest | Ref.  1.05 (0.72, 1.55)  **1.63 (1.11, 2.40)**  1.28 (0.87, 1.88)  **1.89 (1.28, 2.80)** | Ref.  0.82 (0.51, 1.31)  1.22 (0.73, 2.02)  0.89 (0.53, 1.52)  1.31 (0.75, 2.31) |
| More than one child vs one child | 0.98 (0.75, 1.27) | 0.85 (0.65, 1.13) |
| Preterm versus term birth | 0.94 (0.66, 1.34) | 0.93 (0.64, 1.37) |
| Infant breastfed in the last day versus not breastfed | 1.18 (0.92, 1.51) | 1.30 (0.96, 1.77) |

Odds ratio (OR); Confidence Interval (CI).

**^1^** Adjusted for confounders and water, animal, sanitation, hygiene, food, and flooring variables of interest.
